# THE EFFECTIVENESS OF SPIRITUAL SUPPORT FOR CANCER PATIENTS TO IMPROVE THE QUALITY OF LIFE: A LITERATURE REVIEW

**DOI:** 10.1101/2023.10.31.23297645

**Authors:** Ernita Rante Rupang, Moses Glorino Rumambo Pandin, Esti Yunitasari

## Abstract

**Background:** The efficacy of spiritual support for cancer patients is crucial in the course of treatment. As a result, it is crucial to consider the mentoring techniques employed. The purpose of this review of the literature is to methodically look at worldwide research from the previous ten years that has been published in English-language journals from different nations that relates to the usefulness of spiritual support for cancer patients to improve quality of life

**Method:** The method used in this literature review is by database search strategy: data search using a systematic and comprehensive electronic database from several indexing and hand searching. The following indexes were searched for research data: Scopus, CINAHL, PubMed, and Proquest. By utilizing the search terms “OR” spirituality accompanying “AND” quality of life, spirituality support. The eligibility requirements established by the PICO model (Population, Intervention, Comparison/controls, Outcome) were taken into consideration when conducting the article search. Population: cancer patients, Intervention: Spiritual support, Comparison: Intervention provided, Outcome: Improved quality of life.

The process of selecting and extracting data involves stating the problem to be studied, pertinent groups, intervention to be provided, and results to be measured. The author then goes on to clarify the selection and analysis process by outlining the specific inclusion and exclusion criteria as follows: **The inclusion criteria** for the articles reviewed were: the article had to be a full paper, the research subjects were cancer patients, the intervention carried out was spiritual assistance with a comparison of cancer patients without usual care spiritual assistance. An improvement in cancer patients’ quality of life is the search’s result. **The exclusion criteria** for articles reviewed are: articles other than English, research articles before 2017 and articles that are not free access

**Result:** 11 articles were examined. Focuses on providing spiritual support to cancer sufferers, including how to do it and the results. Spiritual support provided through a variety of techniques, including mindfulness, meditation, music therapy, and telephone counseling, encourages patients to be more passionate about their treatments and enhances their quality of life. The patient is religious preferences should be taken into consideration when providing care.

**Conclusion:** Spiritual support has a positive impact on a cancer patient is quality of life by giving them a sense of calm and desire to keep going through the treatment and their daily lives. Through improving coping mechanisms, bringing patients closer to something transcendent (God), and providing a sense of comfort for cancer patients going through chemotherapy, spiritual assistance techniques like mindfulness, listening to music, and counseling can help patients improve their quality of life.

## BACKGROUND

According to WHO (2020), □□cancer is a disease that results in the unchecked growth of an abnormal mass of body tissue cells and can impact the nearby organs. Worldwide, there are a lot more people who are developing cancer□□. According to statistics from around the world, there are 2,261,419 breast cancer patients (11,7%), 2,206,771 lung cancer patients (11,4%), and 1,931,590 people (10%) with colorectal cancer. 1,414,259 instances (7,3%) of cancer cases were due to prostate cancer, whereas 1,089,103 cases (5,6%) were due to colon cancer, liver 905,667 cases (4,7%), and cervical cancer 604,127 cases (3,1%), followed by 8,879,843 cases (46%) of other cancer kinds.

Based on research conducted globally in 2022, the International Agency for Cancer Patients reports that 9,956,133 people die from cancer worldwide and the greatest, accounting for 1,796,144 fatalities (18%), is lung cancer.. It was followed by colorectal cancer (935,173 deaths), liver cancer (830,181 deaths 8%), colon cancer (768,793 deaths 7%), breast cancer (684,996 deaths 9%), esophageal cancer (544,076 deaths 5%), pancreatic cancer (466,003 deaths 4%), and other cancers (3932,768 deaths 39%) respectively.

Statistics from the Indonesian Ministry of Health (2022) place Southeast Asia eighth globally in terms of new cancer cases per 100,000 people. Among Indonesians, breast cancer is the most common type of cancer. □□In Indonesia, lung cancer accounts for the bulk of cancer cases with 19,4 occurrences per 100,000 individuals and an average death rate of 10,9 cases per 100,000 individuals. The second-highest fatality rate is from liver cancer, with an average of 7.6 cases and 12,4 occurrences per 100,000 individuals. As per the Ministry of Health (2019), breast cancer affects more women than any other disease, with an average mortality rate of 17 per 100,000 people and 42.1 instances per 100,000 educators. The second most frequent cancer in women is breast cancer, which has an average death rate of 13.9 per 100,000 persons and an incidence rate of 23.4 per 100,000□ □.

One of the repercussions of cancer on patients is disruption of many aspects of life, including emotional, psychological, and spiritual activities. (Kiena et al., 2018). This is consistent with other research findings (Majd et al., 2015) that cancer can result in a spiritual crisis that has an impact on all facets of life. According to Izci et al. (2018), cancer patients’ quality of life is impacted. As a result, it is crucial to focus on the ministry of spiritual fulfillment.

According to Ediati et al. (2016), An individual’s spiritual well-being is a result of their surroundings, God, and themselves being in balance. This state is dynamic and includes an element of drive to pursue a life purpose that is ultimately held to be the truth. A person with a fatal condition, in this case a cancer patient, needs to take action by receiving spiritual support, which ultimately enables the patient to discover the highest meaning and purpose in life. A variety of techniques can be used to provide this kind of spiritual support. One of them is mindfulness, which was created by other earlier scholars, including Christian mindfulness (Cernetic et al., 2018). Finding efficient strategies or techniques to assist cancer patients in accepting their circumstances is crucial. Mentoring is concerned with the spiritual side so that it serves as inspiration for those who are suffering to live their next life, despite the odds. This literature review examines spiritual support generally and lists several methods. □This literature review aims to assess the effectiveness of spiritual care given to cancer patients by individuals who share their religious views and that takes those beliefs into consideration□.

## METHOD

The method used in this literature review is to search for relevant articles published in English using four databases, namely: Scopus, PubMed, Proquest and CINAHL. Research to determine the effect of implementing spiritual assistance for cancer patients according to the specified inclusion criteria. The type of research is cross sectional, control trial, experiment, randomized control trial, mixed method, qualitative phenomenology. The main result is the effect of spiritual assistance on cancer sufferers to improve their quality of life. Open access and publications 2017-2023 included in the literature review

## RESULT

Once a focused search using keywords with Boolean operators “AND” and “OR” has been performed, the author discovered 22,023 from four databases: Scopus (532 articles), CINAHL (226 articles), and PubMed (15,052 articles). PICO stands for Population (individuals affected by cancer).), Intervention (help), Comparison and Results (patient clinical status and cancer risk). The study group examined the inclusion criteria and performed Rayyan and manual analyses on ten papers.

**Figure 1.**
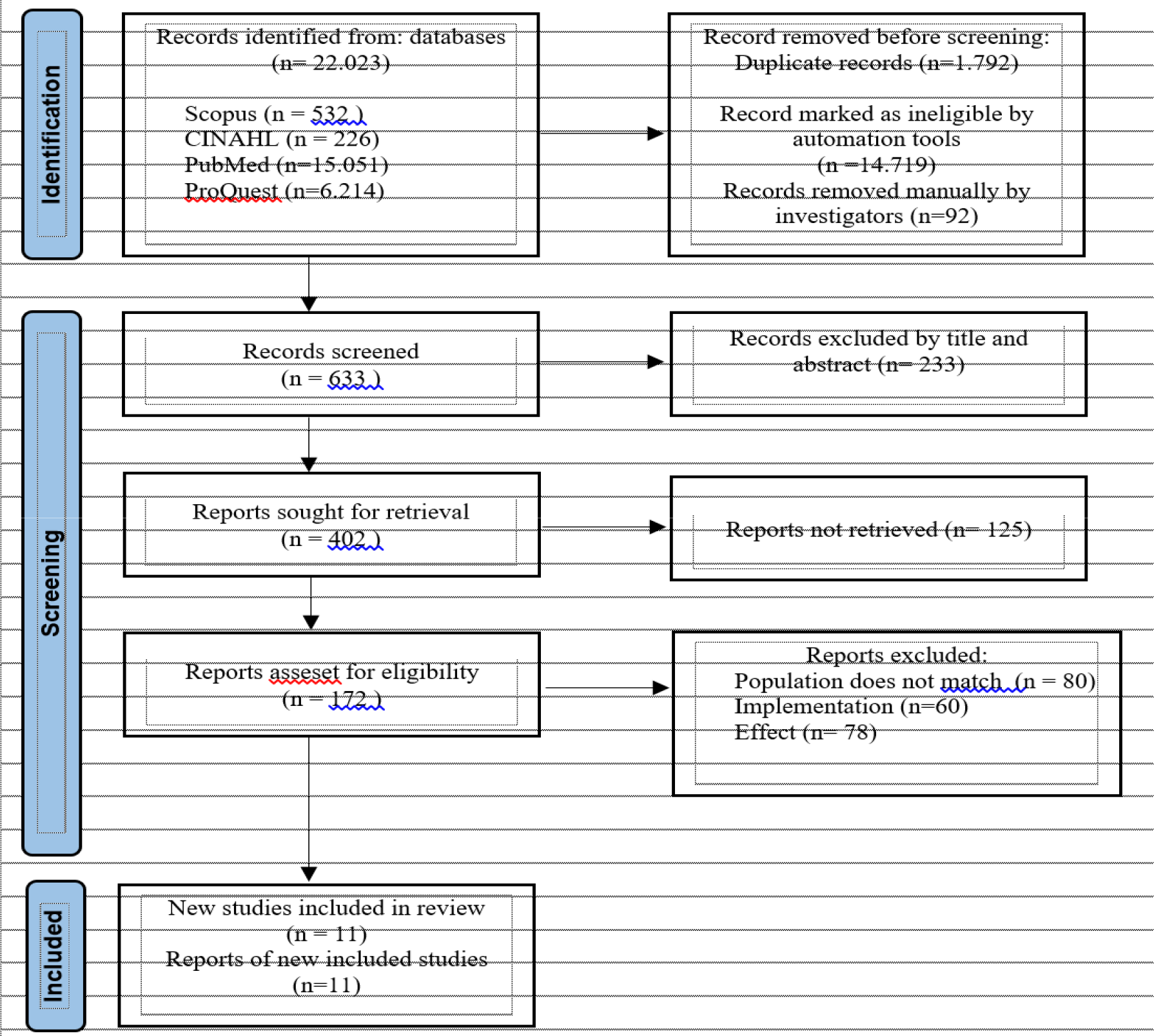
Flowchart for PRISMA.

**Table. 1.** Caracteristics of reviewed studies.

| No | Title, Researcher and Year | Method | Result |
| --- | --- | --- | --- |
| 1 | “Implications for clinical cancer of cancer patients' increased spiritual well-being” (Cheng et al., 2019) | **Design:** Cross sectional<br>**Subjects:** 200 Chinese cancer patients<br>**Variables:** include clinical features, spiritual direction, and religion.<br>**Instrument :** questionnaire<br>**Analysis:** using multiple regression | Finding effective methods to boost spiritual wellbeing is important since it will improve the patient's quality of life |
| 2 | □□A randomized controlled experiment carried out to investigate the effect of spiritual support on the optimism of cancer patients.□□<br>(Afrasiabifar et al., 2021) | **Design:** randomized controlled trial design<br>**Subjects:** 74<br>**Variable:** demographics, chemotherapy treatment times and cancer severity<br>**Instrument:** questionnaire<br>**Analysis:** Examining descriptive data, using chi square and Exact Fisher's, as well as the t test | Psychosocial support enhances cancer sufferers' spiritual health |
| 3 | “A qualitative investigation on Thai breast cancer patients' understanding of spirituality and spiritual well-being”<br>(Phenwan et al., 2019) | **Design:** qualitative study<br>**Subject:** 16 patients<br>**Variable:** age, lifestyle, education, disease stage, length of suffering, and degree of palliative care.<br>**Istrument:** interview guide<br>**Analysis:** descriptif analysis | The survey produced three key themes: a meaningful life, a sense of community, and a closeness to or union with nature. The research recommendation is for future studies to use a secular perspective rather than a religious one, and to be more comprehensive. |
| 4 | “Experience of nasopharyngeal cancer patients following chemoradiation treatment: a phenomenological study”<br>(Sucipto et al., 2019) | **Design:** descriptive phenomenology<br>**Subject:** 11 patient<br>**Variable:** cancer patient treatment, cancer patient experiences, and nasopharyngeal cancer<br>**Instrument:** interview guide<br>**Analysis:** Colaizzi | The study identified three themes: decreased social engagement, support for cancer patients, and xerostomia as the primary physical complaint expressed by patients. |
| 5 | “A pilot study looked at how cancer survivors' spiritual wellness was affected by mindfulness meditation and mild yoga.”<br>(Bryan et al., 2021) | **Design:** mixed method, quasi exsperiment<br>**Subject:** 20 patient<br>**Variable:** physical, spiritual, emotion<br>**Instrument:** questionnaire<br>**Analysis:** t test, thematic inductive | Patients typically see benefits in their spirituality, emotions, ability to manage stress, sleep, and social interactions. |
| 6 | “The effects of church visits on the spiritual welfare and comfort levels of cancer patients”<br>(Watania et al., 2021) | **Design:** cross sectional<br>**Subject:** 146 patient<br>**Variable:** demographics, spiritual well-being, comfort level<br>**Instrument:** questionnaire<br>**Analysis:** bivariate analysis is Chi square and multivariat analysis is logistic regression | The most important aspect for cancer patients is spirituality, especially church attendance, which significantly affects the patient's spiritual well-being. |
| 7 | “Relationship between breast | **Design:** cross sectional | Only schooling has a |
|  | cancer patients' spiritual well-being and degree of exhaustion”<br>(Komariah et al., 2021) | Subject: 112 patient<br>**Variable:** demographics, spiritual well-being, fatigue,<br>**Instrument:** questionnaire<br>**Analysis:** spearman correlation | substantial impact on the spiritual components of demographic data, which has a large impact on fatigue. The link between fatigue and spiritual health is substantial. |
| 8 | □□A randomized clinical experiment to Finding out how spiritual therapy affects cancer patients' spiritual health(Sajadi et al., 2018) | **Design:** randomized clinical trial<br>**Subject:** 42 female cancer patients<br>**Variable:** demography, illness stage, cancer kind, and spiritual health<br>**Instrument:** questionnaire<br>**Analysis:** Use the Kolmogorov-Smirnov test for normally distributed data, and the exact Fisher or chi-square test for comparisons between control groups. | Following the intervention, the study found a significant difference, and it is advised that women with cancer undergo both conventional medical care and spiritual counseling. |
| 9 | “Children who have cancer and their spiritual lives: a study qualitative descriptive in Lithuania”<br>(Juškauskienė et al., 2023) | **Design:** description qualitative<br>**Subject:** 27 cancer patient<br>**Variable:** spiritual life and cancer-affected kids<br>**Instrument:** interview guide<br>**Analysis:** theme analysis | Four themes emerged from the research: acting normally, community, comfort, and relation with God. |
| 10 | “The efficacy of spiritual interventions for promoting spiritual wellbeing and coping in patients diagnosed with gynecological cancer”<br>(Nasution et al., 2020) | **Design:** quasi eksperiment with pre and post control<br>**Subject:** 108 patient<br>**Variable:** demographics, the severity of the cancer<br>**Instrument:** questionnairre Brief COPE and FACIT-Sp | There was a significant change ($p=0.001$) after coping treatment, and a significant change ($p=0.06$) after a spiritual wellness intervention. □ The findings demonstrated that there was a difference between treatments in terms of average coping ($p=0.004$) and spiritual well-being ($p=0.001$). and control groups□. |
| 11 | □□In older breast cancer survivors, personality, coping, and Social support as a means of promoting long-term well-being CALGB protocol 369901 | **Design:** cross sectional<br>**Subject:** 1028 older women pasien<br>**Variable:** Personality traits, coping mechanisms, and social support as markers of long-term | For each Quality of Life measure, Three distinct trajectories were found: "kept high," "phase change," and "lower scoring but |
|  | (Durá-Ferrandis et al., 2017) | quality of life in elderly breast cancer patients<br>**Instrument:** questionnaire QoL<br>**Analysis:** Chi square tests untuk analisa bivariate dan regresi logistic untuk data multivariat | parallel to the corresponding faster decrease in emotional, physical, and cognitive performance in the elderly (6.9%, 31.8%, and 7.6% of the population, respectively). The "maintained high" group). Lower optimism was only related to poorer emotional functioning, and coping with adaptive mal For all quality of life categories, the adjusted chances of belonging to the group suffering an accelerated deterioration were higher among those with less social support and adjustment. Chemotherapy is connected to cognitive and physical but not emotional trajectories. |

## DISCUSSION

### Spiritual accompaniment

Given that cancer has a high incidence rate, it is currently quite evident how vital spiritual support is for the health industry, especially in the treatment of cancer patients.. (Lebowa et al., 2023). Health professionals must question patients how they apply or experience spiritual or religious topics because the majority of religions hold the belief that life is sacred and must therefore be preserved or extended (Guidozzi & Guidozzi, 2022).

Pastoral care is a crucial component of holistic care, according to (Liu et al., 2023) since it improves the wellbeing of children with cancer. In the implementation of services that focus on patients and family, the spiritual component is acknowledged as a crucial component of palliative care, particularly for cancer patients (National Concensus Project for Quality Palliative Care, 2018). Communication skills are necessary for effective spiritual care in order to comprehend the patient’s spiritual situation. Even when the service provider has provided excellent spiritual care, patients frequently report feeling as though they are not receiving adequate spiritual care (Ellington et al., 2017).

Acting normally, community, comfort, and contact with God are the four aspects that influence the spiritual assistance of cancer patients (Juškauskienė et al., 2023).

### Implementation of accompaniment

As a result of its enormous impact on cancer patients, spiritual support is crucial in patient care and services (Farahani et al., 2022). (Since ancient times, music has been acknowledged as a form of therapy that can help with both physical and mental recovery and treatment (Alvarenga et al., 2018). Cancer patients and those receiving palliative care benefit from music therapy because it lowers pain thresholds, reduce anxiety, and comforts them. As a result, patients who get music therapy utilizing the teleheath music therapy method have a spiritual well-being effect that helps them cope with stress and challenges more calmly (Folsom et al., 2021).

Since social media has a big impact on the treatment process, especially when it comes to improving quality of life, one way to achieve the goal of enhancing support for cancer patients is to improve social support through social media platforms. (Hale et al., 2020). □Specifically, mindfulness meditation enhances cancer patients quality of life□. through encouraging mental acuity, physical health, and well-being (Cheng et al., 2019). The QoL of cancer patients is improved by receiving spiritual support through education (Li et al., 2019).

According to (Phenwan et al., 2019), relationships with family, friends, and the environment all have an impact on one’s spiritual well-being. Cancer patients’ spiritual well-being will increase with improved relationships. According to (Sucipto et al., 2019), adopting spiritual help needs family and social support. This translates to a relationship between all aspects of life. According to (Sajadi et al., 2018), For cancer patients, spiritual therapy offers a number of advantages, supplementing traditional therapy methods.

### The effect of spiritual accompaniment

The patient’s overall quality of life may improve and their physical discomfort may be reduced with effective spiritual assistance. (Ferrell et al., 2022). □□Spiritual support is a crucial factor that needs to be taken into account in services since communication utilizing the telehealth technique is useful in assisting cancer patients in enhancing their quality of life□□(Meneses et al., 2018). In order the patient’s family to more quietly accept the treatment conditions, pastoral care is provided for cancer patients as well (Rajaei et al., 2022). Prioritizing spiritual accompaniment is crucial because it enhances spiritual welfare (Afrasiabifar et al., 2021).

According to (Bryan et al., 2021), mindfullness promotes spiritual wellbeing in cancer patients. The application of religious assistance plays a role in enhancing spiritual welfare, thus it becomes the primary focus in helping cancer patients, according to research (Watania et al., 2021), which supports this. According to (Komariah et al., 2021), spiritual support significantly affects other elements of cancer patients’ lives. According to (Yang et al., 2023) spiritual support provides cancer patients with encouragement and lowers their levels of worry and depression.

### Improvement QoL

The symptoms and function, but not worse than the state of patients who maintain health, according to the EORTC QLQ-C30 results in some scenarios. (Finck et al., 2018). Because of spiritual support, there is a favorable correlation between gratitude and life fulfillment. Patients usually ask their doctors, friends, and family for social support. (Finck et al., 2018). The findings imply that optimism aids in the condition’s management. A more in-depth analysis of the symptoms and functional domains must precede any general evaluation of overall quality of life.

Three trajectories for each Quality of Life domain were identified, supporting (Finck et al., 2018): 31.8%, 6.9%, and 7.6% of The elder group showed maintained high, phase shift (lower but parallel scores to the maintained high group), and faster decrease in terms of emotional, physical, and cognitive function, respectively. Reduced emotional functioning, adaptive maladjustment coping, and low social support were the only factors linked to lower optimism. These traits significantly raise the patient is likelihood of falling into the category of people whose quality of life is fast declining. According to (Durá-Ferrandis et al., 2017), Chemotherapy is associated with changes in cognitive and physical abilities, but not with emotions.

## CONCLUSION

The impact that spiritual support has on cancer patients’ quality of life includes the following: it gives patients a sense of calm so that they can face life and the treatment process with optimism; it gives cancer patients who are undergoing treatment motivation and zeal. Meditation, music listening, and therapy are spiritual support techniques. For cancer patients who are receiving chemotherapy, counseling support might help them feel more at ease. Spiritual accompaniment enables patients to live healthier lives by facilitating improvements in coping mechanisms and a stronger relationship with something transcendent (God). It is crucial to remember that while employing spiritual support techniques to provide spiritual support to cancer patients can improve their QoL, there hasn’t been much research from the reviewed articles on doing so from the perspective of the patient’s chosen religion.

## Data Availability

All data produced in the present work are contained in the manuscript

## ACKNOWLEGGMENTS

That you to the nursing lectures at Airlangga University and who have helped write this article

